# Impact of COVID-19 on the mortality rates for the resident population of the Umbria region in Italy

**DOI:** 10.1101/2020.09.24.20200667

**Authors:** Carla Bietta, Mattia Morini, Asiya Kamber Zaidi, Francesco Cozzolino, Puya Dehgani-Mobaraki

## Abstract

The mortality figures related to the coronavirus pandemic has been the topic of debate lately. Several hypothesis are made regarding the expected number of deaths in a region but there are various factors governing the same. In this paper, we have discussed the mortality figures in the Umbria region after analyzing the data from the national Health registry between December 2019 to April 2020; the period of infection and its comparison with the data from previous five years. The factors governing these figures were studied including temperature, standard mortality rates, territorial distribution, death due to all cases as well as the non-COVID deaths. A sharp increase in mortality figures was observed for the month of march and low temperature also had a role to play. However the difference when compared to previous 5 years was not significant as was expected at the start of the study. A single factor cannot be responsible for the total mortality figures in a region as is frequently predicted.

## Introduction

One of the most debated aspects of this pandemic period has been the mortality due to SARS-CoV-2 infection. There has been a lot of discussion about the actual number of deceased, considering underestimation of reporting, as all deaths has not been RT-PCR tested for Covid-19. This is an indicator influenced not only by the methods of classification of causes of death, but also by the availability of tests to detect the novel coronavirus. *[1]*

This pandemic might have several repercussions, both positive and negative, on the mortality figures. On one hand, we can hypothesize an increase in deaths due to cases with COVID-19, cases with symptoms of COVID-19 that have not been tested and the cases leading to death due to other pathologies (areas with poor access to healthcare). On the other hand, there might also be a reduction in number of deaths from road traffic accidents owing to the lockdown. *[2]*

## Methods

We have discussed the mortality rates of the Umbria Region based on the Regional Health Registry data from December 2019-April 2020(the period hypothetically linked with the spread of the virus) and comparing it with the previous five years(2015-2019). The impact of the pandemic on the mortality figures of the population can be explained by comparing the deaths due to all causes, during the same period, with the average deaths during the previous five years assuming that the spread of the infection could, due to its complexity, produce an increase in deaths that may not be directly attributable to COVID-19 positive deaths.

This information partially manages to fill a heavy information gap in reading the Umbria mortality data, considering the fact that to date the prevention departments do not have a regularly computerized mortality data, named Register of Causes of Death.

## Results

There was a reduction in the mortality figures for the months of December, January, February and April respectively of 11%, 13%, 8% and 6% and an increase of 6% in March. An increase in the number of deaths was observed in months with lower temperatures; starting from the month of March. The standardized mortality rate for the month of March was significantly higher than in February and April for females and in April for males. As per the territorial distribution, the districts of Orvieto and Narni-Amelia experienced a higher mortality rate compared to the other regions. There were no significant difference in the non-COVID-19 related deaths, as compared with the previous five years.

## Discussion

### Comparison of deaths in Umbria Region between 1 December - 30 April, 2015-2020

To compare the mortality, reference was the baseline value determined by an average of daily deaths that occurred between 2015-2019.*This average value was described along with standard deviation, to identify any deviation from the trend (Fig.1a). The 5-day daily average trend of the number of deaths from December 2019-April 2020 remained substantially below the baseline except for the month of March wherein the values exceeded the baseline, even above the upper limit of the standard deviation(Fig.1b).

**Figure 1:**
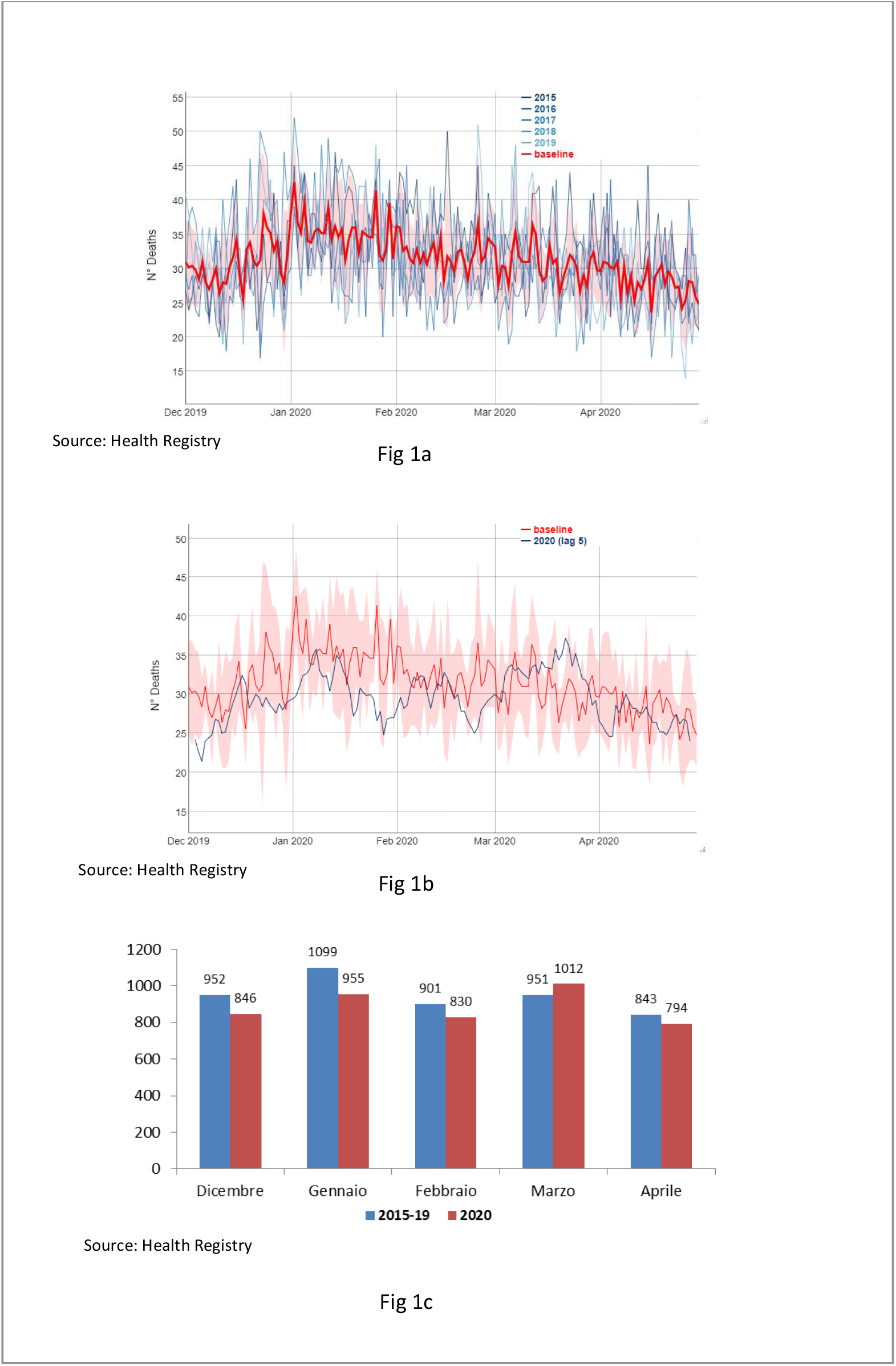
Fig.1a Annual trend and daily average of deaths for all causes. December 1 to April 30. Umbria 2015-19; Fig. 1b The 5-day daily deaths average from all causes from December 1st - April 30th in Umbria. Comparison 2020 and baseline 2015-2019 and Fig.1c. Monthly deaths for all causes from December to April. Umbria. Comparison 2020 and average years 2015-19.

The same comparison with the monthly average of deaths that occurred in the five-year period 2015-19 confirms what has been observed, showing a reduction for the months of December, January, February and April respectively of 11%, 13%, 8% and 6% and an increase of 6% in March(Fig. 1c).

### The influence of temperature

One of the factors that should be considered while comparing mortality between infra-annual periods is the seasonal trend of deaths, for which weather acts as one of the elements determining these oscillations. This influence can be demonstrated by observing the relationship between the average daily temperature and daily mortality for the same period. *[3]* From the available data(Fig. 2a) an increase in the number of deaths was observed in months with lower temperatures; starting from the month of March. Every year, a progressive decrease in the number of deaths was observed corresponding to the rise in average temperatures every day. †For the month of March 2020, on the other hand, there was an increase in the number of deaths despite the rise in temperature(Fig. 2b). This trend therefore shows a direct causal effect of the epidemic in the regional territory.

**Figure 2:**
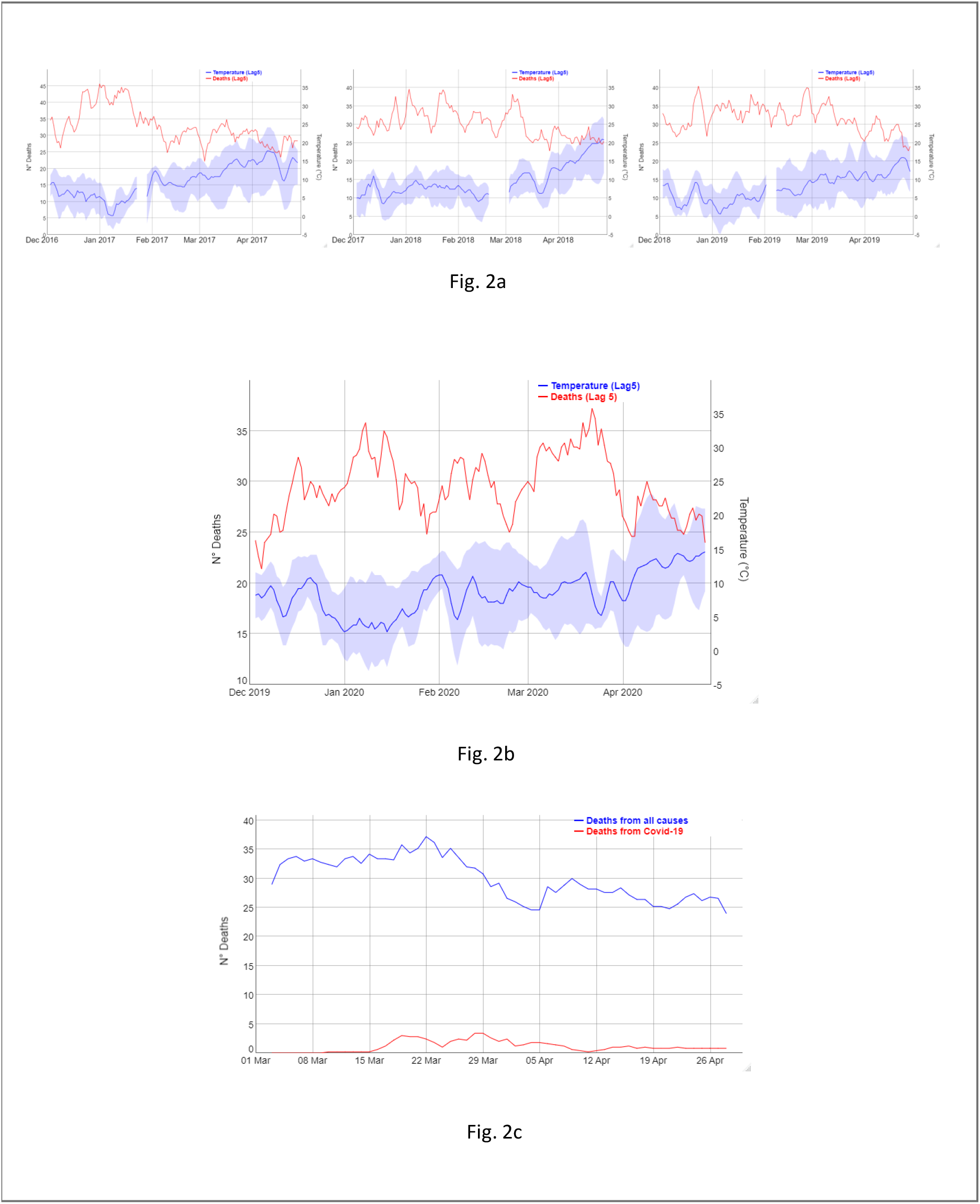
Fig.2a. Trend of the 5-day daily average of deaths for all causes and the 5-day average of average T ° C (minimum and maximum) 1 Dec - 30 Apr. Umbria. Years 2017,2018 and 2019; Fig.2b. Trend of the 5-day daily average of deaths for all causes and the 5-day average of average T ° C (minimum and maximum) 1 Dec - 30 Apr. Umbria. Years 2017, 2018 and 2019 and Fig.2c. Daily average of 5 days of deaths for all causes and for COVID-19 March 1st - April 30th 2020. Umbria.

### The comparison between deaths for all causes and for COVID-19

Umbria region encountered the pandemic a little later compared to other regions, with a few weeks of delay. i.e, the first group of cases were detected at the end of February. This further lead to a delay in the recording of deaths, i.e starting from mid-March. The comparison of average daily deaths to 5 days for COVID-19 for all causes during March-April 2020 showed no significant correspondence in the performance of the two curves(Figure 2c). However, it should be remembered that deaths from COVID-19 are a limited number (N = 67).

### The Comparison of standardized mortality rate

In consideration of the fact that in March there is a greater mortality from all causes compared to the previous five years, in order to verify the hypothesis that even in our region the number of deaths linked to COVID-19 infection may be underestimated, the standardized monthly mortality rate for all causes was calculated for the two periods under review.

The comparison between the standardized mortality rates in March does not show any difference for both sexes between 2020 and the average for the five-year period 2015-19. In comparison between the two periods, however, a lower standardized mortality rate was observed in January 2020. On the other hand, in the comparison between the five months of 2019-20, we observed for the month of March a standardized mortality rate that is significantly higher than in February and April for females and in April for males (Tab.1).

### Territorial distribution

A crude mortality rate was calculated for all causes based on territorial distribution in the first quarter(January-April 2020).From the map it can be inferred that the districts of Orvieto and Narni-Amelia had a higher rate compared to the others.(Fig.3-Tab.2)The comparison with the previous five years shows a higher mortality in 2020 for the same districts than the average of the previous years, highlighting an increase in deaths for these territories compared to the expected 4.4×10,000 inhabitants. The increase in deaths persists even by subtracting the number of deaths attributed to COVID-19 from the total for the four-month period(Table 2).

**Figure 3:**
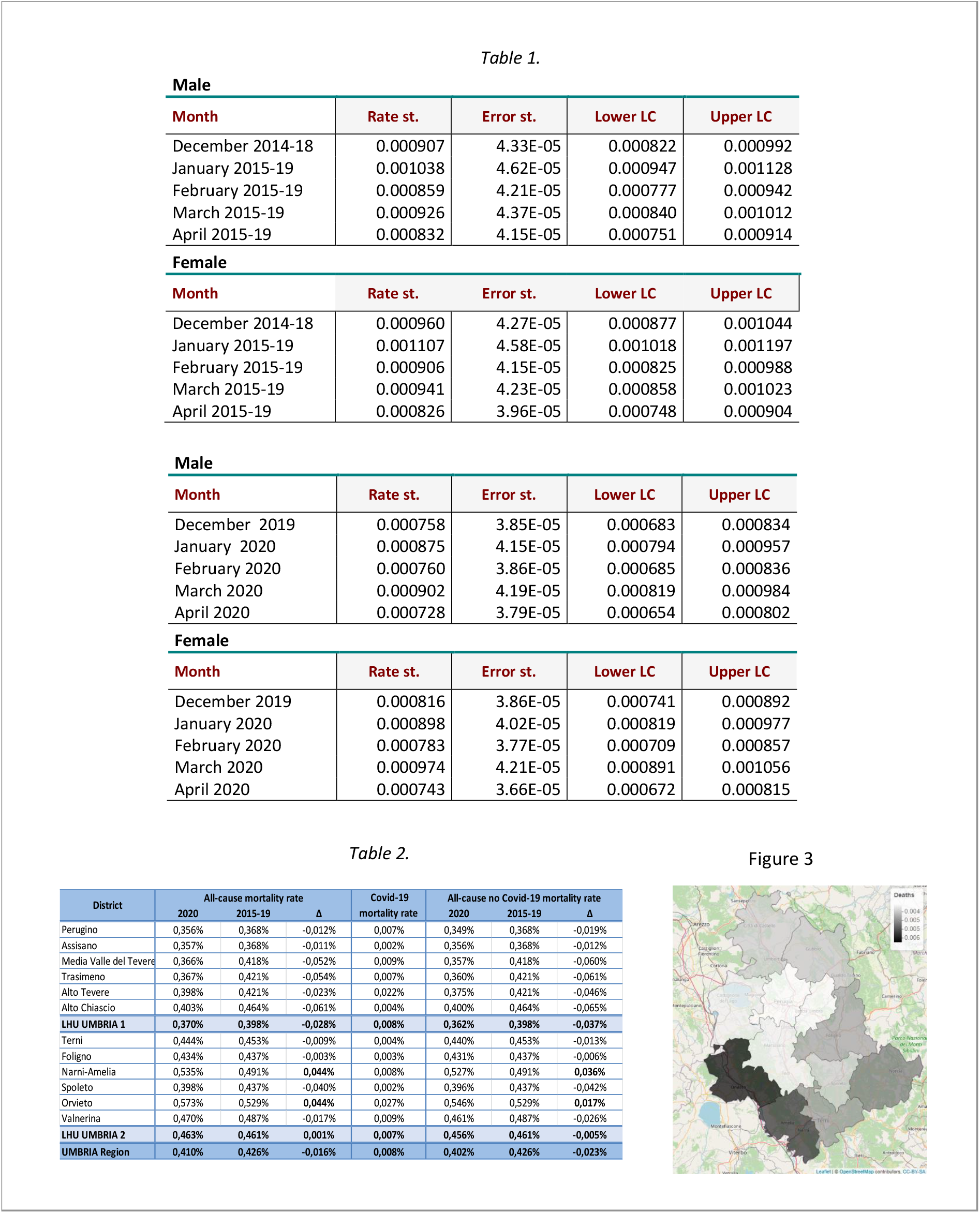
The Standardized mortality rate per month in Table 1; The crude mortality rate by district and Local Health Unit, four months January-April in Table 2; and Crude mortality rate by district. Umbria. Quarter January-April 2020 in the figure.

This could in part support the hypothesis that in these two districts the number of deaths linked to COVID-19 infection may have been underestimated.

### Unintended Mortality

”Unintended” mortality *[4]* due to road accidents and occupational accidents during the lockdown period, 11 March-30 April, was considered to affect the total number of deaths occurring due to all causes. For this purpose, the number of deaths from the aforementioned lockdown cases was estimated(50 days), based on the mortality due to cause information available from *ISTAT* (The National Statistical Institute is an Italian public research body that deals with general censuses of the population, services and industry at national level) and fatal accidents from *INAIL* (The National Institute for Insurance against Accidents at Work is an Italian non-economic public body, subject to the supervision of the Italian Ministry of Labor and Social Policies) in the years 2015-17. This number was then added to the total number of deaths that occurred from 11 March-30 April 2020 and compared with the average deaths for the five-year period 2015-19 referring to the same period. *[5]*

> Average N deaths/year for accidents at work and transport s accidents (years 2015-17) = 75
>
> Estimate N deaths / day = 0.205
>
> Estimated deaths not occurred in the Lockdown = 0.205 x 50gg = 10.3 N deaths from 11 March to 30 April 2020 = 1494
>
> Average N deaths from 11 March to 30 April 2015-19 (five years) = 1484
>
> Δ^§^ (2020 – 2015-19) = + 1.35%

Even considering the deaths that did not occur due to COVID-19, there were no significant differences with the previous five years.

## Conclusions

Data analysis from the National Health Registry to reconstruct the trend of mortality from all the causes and all ages in the Umbria region revealed no significant difference for the four-month period between January-April when compared to the five-year period 2015-19 used as a reference. While considering the increase in deaths in March 2020, consistent with the trend of the epidemic in the region, the comparison between the standardized mortality rates does not show significant differences for both the sexes as compared to the five-year reference period. The territorial distribution of deaths showed a higher mortality rate for the districts of Orvieto and Narni-Amelia. By restricting the comparison to the lockdown period and also taking into account the estimated mortality “not occurring” due to road traffic accidents and fatal accidents at work in the same period, no differences were observed with the previous five years. From the analyzed data, therefore, no elements emerge that suggest a further impact of the COVID-19 pandemic on the total mortality of the population residing in Umbria.

## Data Availability

Can be made available on request.

## Summary Box

### What is already known on this topic?

The pandemic of COVID-19 has caused a major havoc all over the world. It was hypothesized that the disease and the lockdown might have a significant effect on the mortality figures.

### What is added by this report?

The total mortality in a region depends on several factors including the number of test performed, availability and resources, the reporting of data, temperature, and the non-COVID related mortality.

### What are the implications for public health practice?

A single factor cannot be responsible for the total mortality figures in a region as is frequently predicted. When the data is analyzed and compared over 5 years, there was no significant difference in the mortality.

